# Mediation analysis to inform policy on coronary revascularization by expected time to treatment: Analytical framework

**DOI:** 10.1101/2021.12.30.21268556

**Authors:** Boris Sobolev, Lisa Kuramoto

## Abstract

**Objectives:** Clinical guidelines favour coronary artery bypass grafting (CABG) over percutaneous coronary intervention (PCI) for patients with stable complex coronary disease. Yet the benefit of CABG as established in trials may not be generalizable to populations in which treatment method determines time to treatment, typically being longer for CABG. For cases in which the cardiac anatomy is suitable for either treatment, it is unclear whether it is appropriate to recommend CABG, which is likely to be delayed, if PCI can be performed sooner. This paper outlines an analytical framework for a policy analysis of the timing of coronary revascularization.

**Methods:** We constructed a thought experiment to examine whether time to treatment will influence the advantage of CABG. We substantiated the use of mediation analysis to estimate the extent to which differences in outcomes between CABG and PCI would change if times to CABG were the same as times to PCI.

**Results:** We designed a study that uses data from a population-based patient registry to obtain effect measures of mediation analysis: the total effect, the natural indirect effect, and the natural direct effect. The partitioning of the total effect will allow us to estimate the proportional reduction in the risk of an outcome if the time to CABG was similar to that of PCI.

**Interpretation:** Treatment recommendation, resource allocation and scheduling benchmarks will be guided by understanding the extent to which the time to treatment mediates the relation between revascularization method and outcome.

## INTRODUCTION

The narrowing of the coronary arteries that limits blood flow to the heart muscle constitutes a medical condition called coronary artery disease (CAD). The restoration of blood flow, or revascularization, is achieved by either widening or bypassing the narrowing. These two procedures are called percutaneous coronary intervention (PCI) and coronary artery bypass grafting (CABG), respectively. Underlying causes of CAD can lead to another narrowing of the same or a different artery after initial revascularization. Further revascularization is needed when a clinically significant narrowing re-occurs.

There has been debate about the appropriate procedure for patients with multi-vessel or left main disease who do not need emergency treatment.^1-4^ For patients with the cardiac anatomy suitable for either procedure, recent clinical guidelines favour CABG over PCI.^5-8^ This recommendation reflects findings from randomized trials showing that CABG results in higher or similar survival, fewer heart attacks and reduced need for repeat revascularizations, relative to PCI.^9,10^ However, these findings may not be generalizable to populations in which time to treatment depends on the revascularization method, with the wait typically being longer for CABG than PCI. First, the therapeutic effect of revascularization may vary with time. Second, factors that cause variation in time to treatment may also cause the comparative effectiveness of the two treatments to vary from one patient to another. Yet the clinical guidelines do not address the relative benefits of CABG when treatment is delayed.^5,8^ In fact, the effect of CABG versus PCI has never been studied in relation to treatment timing.

This article concerns treatment choice according to expected time to treatment, an issue of substantial interest in the policy-making setting. Imagine a policy maker who considers resource allocation when choosing CABG means a longer wait for treatment than when choosing PCI. The natural question is whether patients will have better outcomes if they undergo PCI instead of CABG, given the uncertain delay for CABG. In the following sections, we describe an analytical framework for examining the effect of changes in CABG treatment time on differences between CABG and PCI outcomes. These new evidence will make it easier to determine the appropriate care for individual patients and guide decisions about resource allocation.

## ANALYTICAL FRAMEWORK

### Appropriateness of timing

For cases in which the cardiac anatomy is suitable for either treatment, it is unclear whether it is appropriate to recommend CABG, which is likely to be delayed, if PCI can be performed sooner. Traditionally, appropriate care has been linked to the benefits and risks of the revascularization procedure itself.^7^ Some have hypothesized that CABG have better capacity for preventing cardiac events after initial revascularization on the premise that PCI targets flow-limiting narrowing, whereas CABG restores flow beyond the narrowing.^3^

Our concern here is the benefits and risks associated with the time to treatment. We observe that a choice of the revascularization method will influence when the patient undergoes treatment, a *mediating* factor that affects outcomes by prolonging the patient’s exposure to narrowing in the coronary artery (Figure 1). Disease progression during an extended treatment delay is thought to increase disease severity and lead to incomplete revascularization.^11,12^ For coronary revascularization, therefore, the appropriateness of care should address the concern that some patients may not receive treatment when they would benefit most.^13^

**Fig 1.**
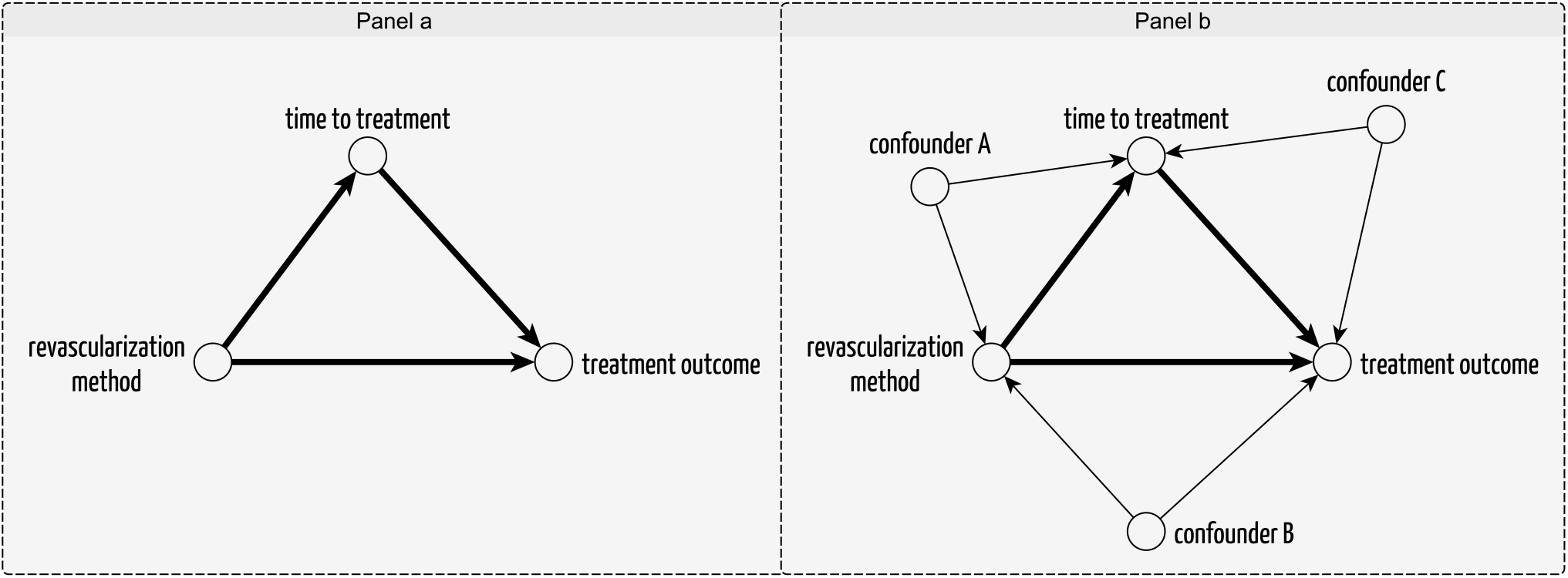
Basic mediation diagram, Panel A: The total effect of revascularization method on treatment outcome is produced through direct influence and through influence on timing of treatment, which in turn may influence outcomes. The revascularization methods have differential capacity for preventing cardiac events, and the choice of revascularization method determines the time to treatment, which is typically longer for CABG than for PCI; extended delays to initial treatment increase disease severity and lead to incomplete revascularization. Confounded mediation diagram, Panel B: Confounders for treatment-outcome, treatment-timing, and timing-outcome associations.

### Thought experiment

Our goal is to assess the difference in outcomes while making each patient (whether undergoing CABG or PCI) wait the same time as if they were undergoing PCI. Determining this difference would require testing the same patient population twice, once with each method, yet doing so would be impossible in the randomized trial setting. Indeed, even if delaying treatment were a true intervention, replacing CABG-induced times to treatment with PCI-induced times would not be equivalent to conducting a randomized experiment in which the intervention arms had fixed times to treatment.^14^ To determine how long a CABG patient would wait if treated with PCI instead, that patient must first be treated with PCI, and would then no longer be eligible for the CABG arm. However, we can speculate what would happen if CABG were performed within times typical for PCI. Such subjunctive reasoning about a hypothetical situation constitutes a thought experiment. Here, we consider three hypothetical situations (labeled A, B, and C in Box 1) and speculate whether the relative benefits of CABG would change if situations A and C occur instead of situations A and B.

### Mediation analysis

We propose to use mediation analysis to investigate how time to treatment might affect differences in outcomes between CABG and PCI.^15^ The purpose of mediation analysis is to partition the total effect of a treatment choice into the effect due to solely the treatment method (direct effect) and the effect due to treatment-induced timing (indirect effect).^16^ This partitioning will allow us to estimate the portion of the total effect attributable to the time to revascularization. This measure shows what would happen to the effect of choosing CABG if policy makers were to remove its influence on time to treatment.

First, we seek to determine the outcomes that would be expected if all patients were to undergo PCI and if all patients were to undergo CABG, within the times naturally occurring for each method. Then we pose a question: “What would be the outcomes of choosing CABG instead of PCI if the choice of treatment method **had no influence** on the time to treatment?” Here, we are seeking to determine the difference in outcomes that would be expected if all patients were to undergo PCI and if all patients were to undergo CABG, both within the time to treatment that is typical for PCI.

Fundamental to this reasoning is the concept of the typical, or naturally occurring, time to treatment. We assume that time to treatment varies from one patient to another as governed by factors related and unrelated to revascularization method. We interpret a counterfactual situation in which all factors that determine the time to revascularization with PCI remain in play, to ensure that the time to CABG would be the same as if PCI had been chosen.

## STUDY DESIGN

### Target population

The target population is patients with complex CAD and stable angina that persists despite drug therapy, with cardiac anatomy suitable for PCI or CABG. We define complex disease as significant narrowing (1) in the left main stem, with or without involvement of coronary arteries; (2) in each of the three major coronary arteries (left anterior descending, left circumflex, right coronary); or (3) in two of the three major coronary arteries, with or without involvement of the proximal left anterior descending artery. The time to treatment varies from one patient to another because of factors that may also cause variation in the comparative effectiveness of the two treatments in different patients.

### Observational study

We will use observational data to assess the extent to which times to treatment mediate the comparative effectiveness of the two treatment methods. We plan to use population-based data from a dedicated patient registry that records the times to treatment as they occurred naturally in practice. The databases of the American College of Cardiology Foundation and the Society of Thoracic Surgeons (STS) are just two examples of reputable data sources.^17^

### Study population

Any patient will be eligible for inclusion in our study if the following conditions are met: (1) the patient underwent a first-time non-urgent isolated CABG or single-session PCI procedure for multi-vessel or left main CAD in a recent era (after 2010); (2) the patient did not present with an acute coronary syndrome; (3) clinical and treatment data for the patient were available in a dedicated, population-based registry; and (4) the patient survived the index revascularization. A recent trial showed that only 60% of patients with previously untreated multi-vessel or left main disease were anatomically and clinically suitable for both PCI and CABG.^18^ Of the remaining 40%, some were suitable only for CABG because of high anatomic complexity, and the remainder were suitable only for PCI because of high operative risks. Therefore, excluded from our study population will be patients with complex anatomy, based on the SYNTAX score, and patients with high operative risks, based on the STS score.^7^ Patients undergoing staged PCI will also be excluded, to avoid misclassification with repeat revascularization.

### Interventions

Three hypothetical scenarios related to the method and timing of revascularization define interventions: (1) single-stage PCI with natural PCI timing, (2) isolated CABG with natural CABG timing, and (3) isolated CABG but with natural PCI timing. In the context of policy making, the latter situation can be arranged using appropriate resource allocation and scheduling guidelines. In the thought experiment, we will create such a situation by allowing the time to treatment to take on a natural value for PCI.

### Outcomes

We will use the standard set of long-term outcomes for patients with CAD developed by the International Consortium for Health Outcomes Measurement.^19^ These outcomes include all-cause mortality, heart attack, stroke, heart failure, and repeat revascularization during 5-year period after initial revascularization.

### Mediator

The mediator will be the number of weeks between the date of the treatment decision and the treatment date (i.e., the time to treatment). Our choice of time unit was guided by the rate of residual disease progression, whereby another narrowing of a coronary artery may occur within several weeks of the initial revascularization.^20,21^

### Effect measures

The effect measures of mediation analysis will be obtained using the Neyman model of potential outcomes of hypothetical interventions.^16^ First, we will estimate the total effect of CABG versus PCI as the difference in outcomes if all patients undergo CABG or PCI within the time to treatment that is typical for these procedures (Table 1). Next, we will estimate the natural indirect effect of choosing CABG as the difference in outcomes if all the patients were to undergo CABG within the time to treatment that is typical for CABG and within the time to treatment that is typical for PCI. Then, we will estimate the natural direct effect of choosing CABG as the difference in outcomes if all patients were to undergo CABG and PCI within the time to treatment that is typical for PCI.

**Table 1.**
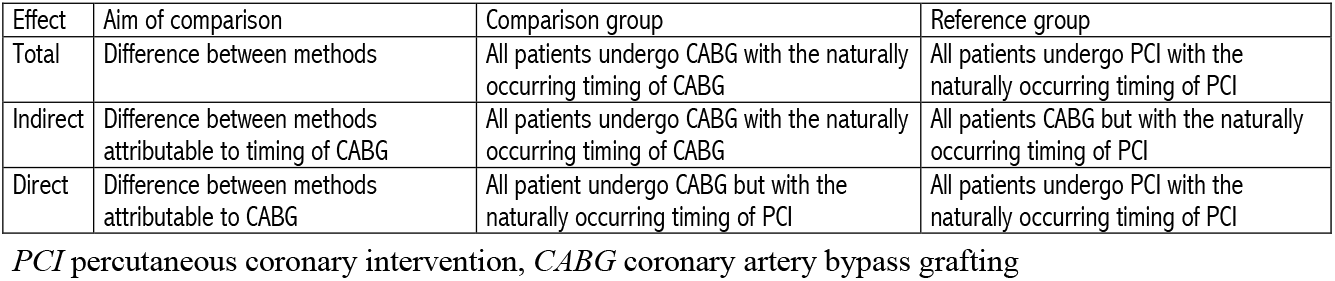
Partitioning the total effect of choosing CABG over PCI into natural direct and indirect effects

| Effect | Aim of comparison | Comparison group | Reference group |
| --- | --- | --- | --- |
| Total | Difference between methods | All patients undergo CABG with the naturally occurring timing of CABG | All patients undergo PCI with the naturally occurring timing of PCI |
| Indirect | Difference between methods attributable to timing of CABG | All patients undergo CABG with the naturally occurring timing of CABG | All patients CABG but with the naturally occurring timing of PCI |
| Direct | Difference between methods attributable to CABG | All patient undergo CABG but with the naturally occurring timing of PCI | All patients undergo PCI with the naturally occurring timing of PCI |

*PCI* percutaneous coronary intervention, *CABG* coronary artery bypass grafting

### Confounders

Causal attribution of variation in outcomes to changes in treatment and mediator could be achieved by conditioning on factors that are sufficient to block all influences that may produce the treatment-outcome, treatment-mediator, and mediator-outcome associations in the absence of causation (Fig. 1). In this context, conditioning refers to stratification according to combinations of factors and examination of the exposure-outcome association at different values of observed times to treatment in every stratum separately. Using a causal diagram,^22^ we will attempt to find a common set of conditioning variables. Should the effect of revascularization method truly vary with timing, we would be able to estimate the natural direct effect by stratifying on confounders of the method-timing association and taking a population average of effects observed within each stratum over observed times to treatment.^23^

## DISCUSSION

Determining the appropriate treatment for patients with stable multi-vessel and left main disease is a complex decision process when the cardiac anatomy is suitable for either CABG or PCI.^1^ Recent guidelines stratify the appropriateness of the revascularization method according to anatomic complexity, risk of postoperative mortality, presence or absence of diabetes, and presence or absence of left main disease.^5,7^ Several authors have argued, however, that other contributing factors, such as local expertise and patient preference, could favour one revascularization method over another.^24^ In health systems where budgetary considerations may delay patients’ planned treatments,^25^ the time to treatment is one factor that has not been studied in the context of comparative treatment effectiveness.^12^ However, there is a strong argument that doctors have a duty to inform their patients of the expected delays and the associated risks when choosing between treatments.^26^

This paper presents a conceptual framework for policy analysis regarding time to treatment in patients undergoing coronary revascularization (Box 2). We argue that the comparative efficacy of the two treatments established in clinical trials may not be applicable to populations in which longer time to CABG reduces its therapeutic effect. We construct a thought experiment to see if time to treatment would affect the benefit of CABG. We justified the use of mediation analysis to assess how much the differences in outcomes would have changed if patients who received CABG had the same treatment time as patients who received PCI.

What is the problem to which mediation analysis is a solution? To properly distinguish between the effect of choosing CABG and the effect of its timing, we need to contrast CABG and PCI outcomes when both treatments are performed within the time typical for PCI. Replacing CABG-induced times with PCI-induced times is not equivalent to setting the times to treatment by means of a randomized experiment.^14,27^ Following Greenland and Robins, we submit that with randomization of patients to various combinations of treatment method and treatment timing, would fail to distinguish the effect of choosing CABG from the effect of treatment timing induced by this choice.^27^ Instead, we will deactivate the influence of the revascularization method on time to treatment through mediation analysis. Using observational data from a population-based patient registry, we are seeking to project treatment outcomes under various scenarios of revascularization method and timing as if they had been implemented for the same patient population.

Our projections will quantify the effect of changes in time to treatment for CABG on differences between CABG and PCI outcomes. We will obtain effect measures of mediation analysis: the total effect, the natural indirect effect, and the natural direct effect. The total effect will show the difference in outcomes between PCI and CABG produced through direct influence of the treatment method and through influencing the time to treatment, which in turn may influence outcomes. The indirect effect will show the difference in outcomes that is attributable to timing. The direct effect will show the difference in outcomes between that is attributable solely to the chosen method. The partitioning of the total effect will further allow us to estimate the proportional reduction in the risk of an outcome if the time to CABG was similar to that of PCI.

## CONCLUSION

The choice of treatment for patients with an indication for coronary revascularization is routinely made without robust evidence about the effect of treatment timing. We offer an analytical framework for estimating the extent to which changes in time to treatment might influence the difference in outcomes between CABG and PCI. The findings on benefits and risks of performing revascularization within a certain time will guide multidisciplinary teams in determining whether PCI or CABG is the appropriate method for individual patients. Resource allocation and scheduling benchmarks will be guided by understanding the extent to which the time to treatment mediates the relation between revascularization method and outcome.

## Data Availability

We constructed a thought experiment to examine whether time to treatment will influence the existing advantage of of one study over another.

## ACKNOWLEDGEMENT

The authors are grateful to Guy Fradet, MD, and Simon Robinson, MD, for useful discussions of ideas presented here; Peggy Robinson, ELS, for presubmission manuscript editing; and to Sean Hardiman for literature search assistance.

## AUTHOR CONTRIBUTIONS

Boris Sobolev and Lisa Kuramoto contributed to the conception and rationale of the study, and to the selection of statistical methodology. Boris Sobolev and Lisa Kuramoto drafted the manuscript and revised it critically for important intellectual content, gave final approval of the version to be published and agreed to be accountable for all aspects of the work.

## Boxes

Box 1. Hypothetical situations considered in the thought experiment

Situation

A: All patients undergo PCI with the naturally occurring timing of PCI

B: All patients undergo CABG with the naturally occurring timing of CABG

C: All patients undergo CABG but with the naturally occurring timing of PCI

*PCI* percutaneous coronary intervention, *CABG* coronary artery bypass grafting

Box 2. Conceptual framework for investigating the effect of changes in the time to treatment on differences in outcomes of revascularization with CABG and PCI in patients with stable angina and complex coronary artery disease

- The premise of appropriate care is timely access to needed treatment.
- CABG is thought to have better capacity for preventing cardiac events after initial revascularization.
- When the cardiac anatomy is suitable for either CABG or PCI, the chosen method determines time to treatment.
- The longer times to treatment for CABG may lessen the relative benefits of this procedure as established in clinical trials.
- The appropriate choice of revascularization method considers effectiveness of CABG versus PCI by treatment timing.
- The policy analysis speculates about outcomes if CABG patients were to have the same time to treatment as PCI patients.
- Randomization by method and timing would fail to distinguish the effect of treatment from the effect of treatment-induced timing.
- Mediation analysis partitions the overall effect of CABG versus PCI into the effects due to treatment and timing.
- The concept of naturally occurring treatment times is fundamental to this mediation analysis.
- The effects are obtained using the potential outcomes approach for hypothetical scenarios of treatment and timing.
- The results of this study will facilitate the determination of appropriate care for individual patients and will guide resource allocation.

*PCI* percutaneous coronary intervention, *CABG* coronary artery bypass grafting

## REFERENCES

1. Windecker S, Neumann FJ, Juni P, Sousa-Uva M, Falk V. Considerations for the choice between coronary artery bypass grafting and percutaneous coronary intervention as revascularization strategies in major categories of patients with stable multivessel coronary artery disease: an accompanying article of the task force of the 2018 ESC/EACTS guidelines on myocardial revascularization. Eur Heart J 2019;40:204–12.

2. Archbold RA. Comparison between National Institute for Health and Care Excellence (NICE) and European Society of Cardiology (ESC) guidelines for the diagnosis and management of stable angina: implications for clinical practice. Open Heart 2016;3:e000406.

3. Doenst T, Haverich A, Serruys P, et al. PCI and CABG for Treating Stable Coronary Artery Disease: JACC Review Topic of the Week. J Am Coll Cardiol 2019;73:964–76.

4. Mulukutla SR, Gleason T, Sharbaugh M, et al. Coronary Bypass Versus Percutaneous Revascularization in Multivessel Coronary Artery Disease. Ann Thorac Surg 2019.

5. Neumann FJ, Sousa-Uva M, Ahlsson A, et al. 2018 ESC/EACTS Guidelines on myocardial revascularization. Eur Heart J 2019;40:87–165.

6. NICE. Stable angina: management (Clinical guideline No. 126). National Institute for Health and Care Excellence; 2011.

7. Patel MR, Calhoon JH, Dehmer GJ, et al. ACC/AATS/AHA/ASE/ASNC/SCAI/SCCT/STS 2017 Appropriate Use Criteria for Coronary Revascularization in Patients With Stable Ischemic Heart Disease: A Report of the American College of Cardiology Appropriate Use Criteria Task Force, American Association for Thoracic Surgery, American Heart Association, American Society of Echocardiography, American Society of Nuclear Cardiology, Society for Cardiovascular Angiography and Interventions, Society of Cardiovascular Computed Tomography, and Society of Thoracic Surgeons. J Am Coll Cardiol 2017;69:2212–41.

8. Teo KK, Cohen E, Buller C, et al. Canadian Cardiovascular Society/Canadian Association of Interventional Cardiology/Canadian Society of Cardiac Surgery position statement on revascularization--multivessel coronary artery disease. Can J Cardiol 2014;30:1482–91.

9. Head SJ, Milojevic M, Daemen J, et al. Mortality after coronary artery bypass grafting versus percutaneous coronary intervention with stenting for coronary artery disease: a pooled analysis of individual patient data. The Lancet 2018;391:939–48.

10. Sipahi I, Akay MH, Dagdelen S, Blitz A, Alhan C. Coronary artery bypass grafting vs percutaneous coronary intervention and long-term mortality and morbidity in multivessel disease: meta-analysis of randomized clinical trials of the arterial grafting and stenting era. JAMA Intern Med 2014;174:223–30.

11. Hambraeus K, Jensevik K, Lagerqvist B, et al. Long-Term Outcome of Incomplete Revascularization After Percutaneous Coronary Intervention in SCAAR (Swedish Coronary Angiography and Angioplasty Registry). JACC Cardiovasc Interv 2016;9:207–15.

12. Head SJ, da Costa BR, Beumer B, et al. Adverse events while awaiting myocardial revascularization: a systematic review and meta-analysis. Eur J Cardiothorac Surg 2017;52:206–17.

13. Graham MM, Knudtson ML, O’Neill BJ, Ross DB. Treating the right patient at the right time: Access to cardiac catheterization, percutaneous coronary intervention and cardiac surgery. Can J Cardiol 2006;22:679–83.

14. Lange T, Hansen KW, Sorensen R, Galatius S. Applied mediation analyses: a review and tutorial. Epidemiol Health 2017;39:e2017035.

15. Lee H, Herbert RD, McAuley JH. Mediation Analysis. JAMA 2019;321:697–8.

16. Pearl J. Interpretation and identification of causal mediation. Psychol Methods 2014;19:459–81.

17. Weintraub WS, Grau-Sepulveda MV, Weiss JM, et al. Comparative effectiveness of revascularization strategies. N Engl J Med 2012;366:1467–76.

18. Head SJ, Holmes DR, Jr., Mack MJ, et al. Risk profile and 3-year outcomes from the SYNTAX percutaneous coronary intervention and coronary artery bypass grafting nested registries. JACC Cardiovasc Interv 2012;5:618–25.

19. McNamara RL, Spatz ES, Kelley TA, et al. Standardized Outcome Measurement for Patients With Coronary Artery Disease: Consensus From the International Consortium for Health Outcomes Measurement (ICHOM). J Am Heart Assoc 2015;4.

20. Cutlip DE, Chauhan MS, Baim DS, et al. Clinical restenosis after coronary stenting: perspectives from multicenter clinical trials. J Am Coll Cardiol 2002;40:2082–9.

21. Pate GE, Lee M, Humphries K, et al. Characterizing the spectrum of in-stent restenosis: implications for contemporary treatment. Can J Cardiol 2006;22:1223–9.

22. Williamson EJ, Aitken Z, Lawrie J, Dharmage SC, Burgess JA, Forbes AB. Introduction to causal diagrams for confounder selection. Respirology 2014;19:303–11.

23. Richiardi L, Bellocco R, Zugna D. Mediation analysis in epidemiology: methods, interpretation and bias. Int J Epidemiol 2013;42:1511–9.

24. Head SJ, Kaul S, Mack MJ, et al. The rationale for Heart Team decision-making for patients with stable, complex coronary artery disease. Eur Heart J 2013;34:2510–8.

25. Sobolev B, Fradet G. Delays for coronary artery bypass surgery: how long is too long? Expert Rev Pharmacoecon Outcomes Res 2008;8:27–32.

26. O’Neill BJ, Brophy JM, Simpson CS, et al. Treating the right patient at the right time: access to care in non-ST segment elevation acute coronary syndromes. Can J Cardiol 2005;21:1149–55.

27. Robins JM, Greenland S. Identifiability and exchangeability for direct and indirect effects. Epidemiology 1992;3:143–55.

